# Integrated Bioinformatics Analysis Identifies and Validates Novel Cellular Senescence-Associated Genes in Sepsis and Sepsis-Induced ARDS

**DOI:** 10.64898/2026.03.30.26349474

**Authors:** Panpan Li, Yongbing Yu, Jihua Feng, Shanshan Huang, Jianfeng Zhang

## Abstract

Sepsis can lead to acute respiratory distress syndrome (ARDS) and is associated with a high mortality rate. This study investigated cellular senescence-related genes in sepsis and sepsis-induced ARDS to identify novel biomarkers. Using bioinformatics analyses including WGCNA and machine learning on public datasets, six hub genes (NFIL3, GARS, PIGM, DHRS4L2, CLIP4, LY86) were identified. These genes showed strong diagnostic value and were associated with immune cell infiltration and key pathways. Validation in lipopolysaccharide (LPS)-stimulated neutrophils showed significant upregulation of NFIL3. The findings highlight the role of cellular senescence in pathogenesis and identify promising therapeutic targets for sepsis-induced ARDS.

## 1. Introduction

Sepsis is a systemic inflammatory response syndrome caused by a dysregulated host response to infection^[1]^, which can lead to multi-organ dysfunction. It affects an estimated 19–48.9 million people worldwide annually and remains a leading cause of death among critically ill patients, despite substantial advances in treatment^[2]^. ARDS, a common and life-threatening complication of sepsis, is characterized by damage to the alveolar-capillary membrane. This injury leads to pulmonary edema and hypoxemia^[3]^. Compared to patients with ARDS not associated with sepsis, those with sepsis-induced ARDS demonstrate higher overall disease severity, delayed recovery from lung injury, and increased mortality^[4]^. Therefore, identifying key molecules in sepsis-induced ARDS and discovering biomarkers for early diagnosis and potential therapeutic targets are critical steps toward reducing its mortality.

Over the past decade, accumulating evidence has established sepsis as a potent stressor capable of inducing cellular senescence^[5–8]^. Cellular senescence is likely to play a significant role in the pathogenesis and long-term outcomes of sepsis and sepsis-induced ARDS. Nevertheless, the molecular profiling of sepsis from a senescence perspective remains poorly defined, and the prognostic relevance of senescence-related genes in these conditions is still not fully understood.

We retrieved and processed sepsis and sepsis-induced ARDS gene expression datasets from the Gene Expression Omnibus (GEO) database, along with cellular senescence-associated genes from public sources. By integrating differential expression analysis, Weighted Gene Co-expression Network Analysis (WGCNA), logistic regression, and machine learning (ML), we identified key hub genes. Subsequent analyses included diagnostic modeling, immune infiltration profiling, consensus clustering, and single-cell RNA sequencing to uncover molecular mechanisms. Functional enrichment analysis revealed associated biological pathways. Experimental validation using Reverse Transcription quantitative Polymerase Chain Reaction (RT-qPCR) in LPS-stimulated neutrophils confirmed hub gene expression. These findings may support novel diagnostic and therapeutic strategies for sepsis and sepsis-induced ARDS.

## 2. Materials and Methods

### 2.1. Data Acquisition and Preprocessing

Genes were screened using the GEO (http://www.ncbi.nlm.nih.gov/geo) database. The reliable expression profiles GSE66890, GSE65682, and GSE167363 are all from Homo sapiens. GSE66890 as training set includes 28 sepsis samples and 29 sepsis-induced ARDS samples, collected on the GPL6244 [HuGene-1_0-st] Affymetrix Human Gene 1.0 ST Array [transcript (gene) version] platform. GSE65682 as validation set contains 802 blood samples, including 760 sepsis and 42 healthy controls, is based on GPL13667[HGU219] Affymetrix Human Genome U219 Array platform. GSE167363 is a total of 12 sets of single-cell data, which Platform is GPL 24676 Illumina NovaSeq 6000, including 2 cases in the control group and 10 cases in the disease group. The probes were transformed to homologous gene symbols using the platform annotation. Genome-wide expression profiles were obtained via the ‘GEOquery’ R package (v2.70.0), with subsequent batch effect adjustment performed using the ComBat algorithm in the ‘sva’ package (v3.50.0). Differentially expressed genes (DEGs) were identified using The ‘limma’ R package (v3.58.1), Employing “Cellular Senescence” as the primary search descriptor, The 866 genes were downloaded from the CellAge (https://genomics.senescence.info/cells/)^[9]^, and 4336 genes from GeneCards ((GeneCards Version 5.24 (Updated: Mar 28, 2025);https://www.genecards.org/)^[10]^ databases, we final got 4482 Cellular Senescence-associated genes by combining the above genes to remove the weight. The 268 Mitochondrial Energy Metabolism-associated genes were obtained in GeneCards database by using the term “Mitochondrial Energy Metabolism” as the search keyword.

### 2.2. Phenotype scoring of Cellular Senescence

Phenotype scores for Cellular Senescence and Mitochondrial Energy Metabolism were calculated using the z-score algorithm in the GSVA package in R software. Associations in phenotype scores between cellular senescence and Mitochondrial Energy Metabolism were assessed.

### 2.3. Weighted gene co-expression network analysis

WGCNA was implemented to delineate gene modules co-expressed with Cellular Senescence. After filtering low-variance genes, phenotype scores for Cellular Senescence and Mitochondrial Energy Metabolism served as input traits for network construction^[11]^.The optimal soft-thresholding power (β) was determined by evaluating scale-free topology fit indices and mean connectivity across candidate parameters. Using this β-value, a scale-free topology network was generated by converting pairwise spearman correlation coefficients into an adjacency matrix, followed by transformation into a topological overlap matrix (TOM). Gene dissimilarity was quantified as (1 - TOM). For module detection, average linkage hierarchical clustering was performed on the TOM-based dissimilarity matrix, coupled with dynamic tree-cutting to define modules with a minimum size threshold of 100 genes. Finally, module membership (MM) and gene significance (GS) were computed to prioritize key modules.

To identify sepsis and sepsis-induced ARDS associated Cellular Senescence-DEGs (CS-DEGs), We conducted integrative intersection analyses between differentially expressed genes (DEGs) and key module genes across two datasets (GSE66890 and GSE65682).

### 2.4. Logistic Regression Analysis and Machine Learning Analysis

First, we performed logistic regression analysis based on the GSE66890 CS-DEGs and GSE66890 training set. When the dependent variable was binary, and the groups were named as Sepsis-Induced ARDS and Sepsis, we applied logistic regression to analyze the association between independent variables and the dependent variable using P.Value < 0.05 as the criterion to screen CS-DEGs. Next, based on the CS-DEGs included in the logistic regression analysis, we used six common ML algorithms to screen the CS-DEGs: random forest (RF), Bayesian analysis, the Wrapper feature selection (Boruta) method, least absolute shrinkage and selection operator (LASSO), and learning vector quantization (LVQ). 7 candidate genes were consistently prioritized by six distinct machine learning algorithms. Subsequent intersection analysis of these candidates with CS-DEGs from the GSE65682 dataset yielded 6 consensus genes, which were selected as hub genes.

### 2.5. Validation of Hub genes and construction of a nomogram

The expression levels of hub genes were extracted from the GSE66890 and GSE65682 datasets. Wilcoxon rank-sum tests were performed to compare expression differences between disease and control groups, and boxplots were generated to visually present the significantly differentially expressed hub genes. We constructed nomograms to evaluate the diagnostic values of the hub genes using the “rms” package in R. The receiver operating characteristic (ROC) were then used to assess the reliability of the model predictions.

### 2.6. Immune Infiltration Analysis

To evaluate the relative abundance of immune cells in sepsis-induced ARDS samples and control samples, immune cell infiltration analysis was performed using the Single Sample Gene Set Enrichment Analysis^[12]^ (GSEA) based on 28 immune cell gene markers^[13]^. The distribution of immune cell components in each sample was visually presented using bar plots, while differences in immune cell infiltration between the two groups were compared using the Student’s t-test (t-test) and displayed as boxplots. Additionally, to explore the relationship between hub genes and immune cell infiltration, Spearman correlation coefficients were calculated, and interactions hub genes and immune cell were illustrated through correlation heatmaps. Finally based on GSE66890 training set, we selected the immune cells that had the strongest correlation with the single gene and constructed the scatter plots to illustrate the correlation.

### 2.7. Identification of Cellular Senescence subtypes associated with Sepsis-Induced ARDS

To explore potential subtypes of Cellular Senescence in Sepsis-Induced ARDS patients, unsupervised consensus clustering was performed based on the expression profiles of hub genes. We applied “ConsensusClusterPlus” R package to determine the optimal k value for the Cellular Senescence-related clusters^[14]^. The parameters were setted as follows: a maximum of 5 clusters (maxK = 5), 100 iterations, Km clustering algorithm, and Pearson distance. Based on the clustering results, Sepsis-Induced ARDS samples in GSE66890 dataset were categorized into two Cellular Senescence subtypes, labeled as Cluster1 and Cluster2.In addition, GSEA was performed next to analyze the biological functions of two subtypes, the reference gene set “ c2.all.v2024.1.Hs.symbols.gmt” was downloaded from the MSigDB database^[15]^.

### 2.8. Single cell analysis

Cell clustering and identification analysis was performed based on the GSE167363 dataset. Firstly, the expression profile was read in by Seurat package, Quality control (5000>nCount_RNA>200, nFeature_RNA > 200, log10GenesPerUMI> 0.8, mitoRatio > 0.8), and screened out low-expressed genes were performedc, subsequently, we constructed doublet removal using DoubletFinder approach, furthermore, the data were processed by normalization, homogenization and PCA analysis in turn. After data preprocessing, the top 30 principal components were chosen for clustering using FindNeighbors and Findclusters functions set to dims = 1: 30, resolution = 0.8, and Uniform Manifold approximation and Projection (UMAP) was used for cell category visualization. The clusters were annotated by the annotation file that comes with the celldex package and were separately annotated to some immune cells that have important relationships with sepsis occurrence. Finally, marker genes were extracted for each cell subtype from the single cell expression profile by setting the logfc.threshold parameter of FindAllMarkers to 0.5. The genes with |avg_log2FC| > 0.5 and p_val_adj <0.05 were screened as the specific marker genes in each cell subtype. We calculated hub genes expressions in varying classes of cells to display their relationship with cell classes, and screened out the cell classes which was the higher expression of prognostic genes.

### 2.9. Cell culture and RT-qPCR

The human promyelocytic leukemia cell line HL-60 was obtained from the laboratory of Basic Medical Sciences, Guangxi Medical University. The cell line HL-60 was cultured in Iscove’s Modified Dulbecco’s Medium (IMDM) supplemented with 10% fetal bovine serum (FBS) and 1% penicillin-streptomycin (P/S) at 37°C in a humidified incubator with 5% CO₂.

To induce differentiation into neutrophil-like cells (dHL-60), cells were treated with 1.25% dimethyl sulfoxide (DMSO) for 5 days. Post-differentiation, dHL-60 cells were divided into two groups: Control, LPS (1 µg/mL LPS for 8 hours, used to establish a sepsis cell model). Cells were collected for subsequent analysis.

Total RNA was extracted from cells using a commercial Total RNA Extraction Kit for Cells/Tissues (NcmSpin Cell/Tissue Total RNA Kit, China). Reverse transcription was carried out using the PrimeScript RT reagent Kit (Takara) on a ProFlex™ PCR System (Thermo Fisher, USA). qPCR amplification was performed with TB Green Premix Ex Taq (Takara) on a Gentier 96E/96R Real-Time PCR System (Xi’an Tianlong, China). The relative expression of target genes was calculated using the 2−ΔΔCt method. The primer sequences used are listed in Table 1.

### 2.10. Statistical analysis

Differences between two groups were assessed using t-test for normally distributed data and Wilcoxon rank-sum tests for non-normally distributed data. Statistical analysis was performed via R software 4.3.3. Differences were considered statistically significant at *p < 0.05, **p < 0.01, and ***p < 0.001.

**Table 1.**
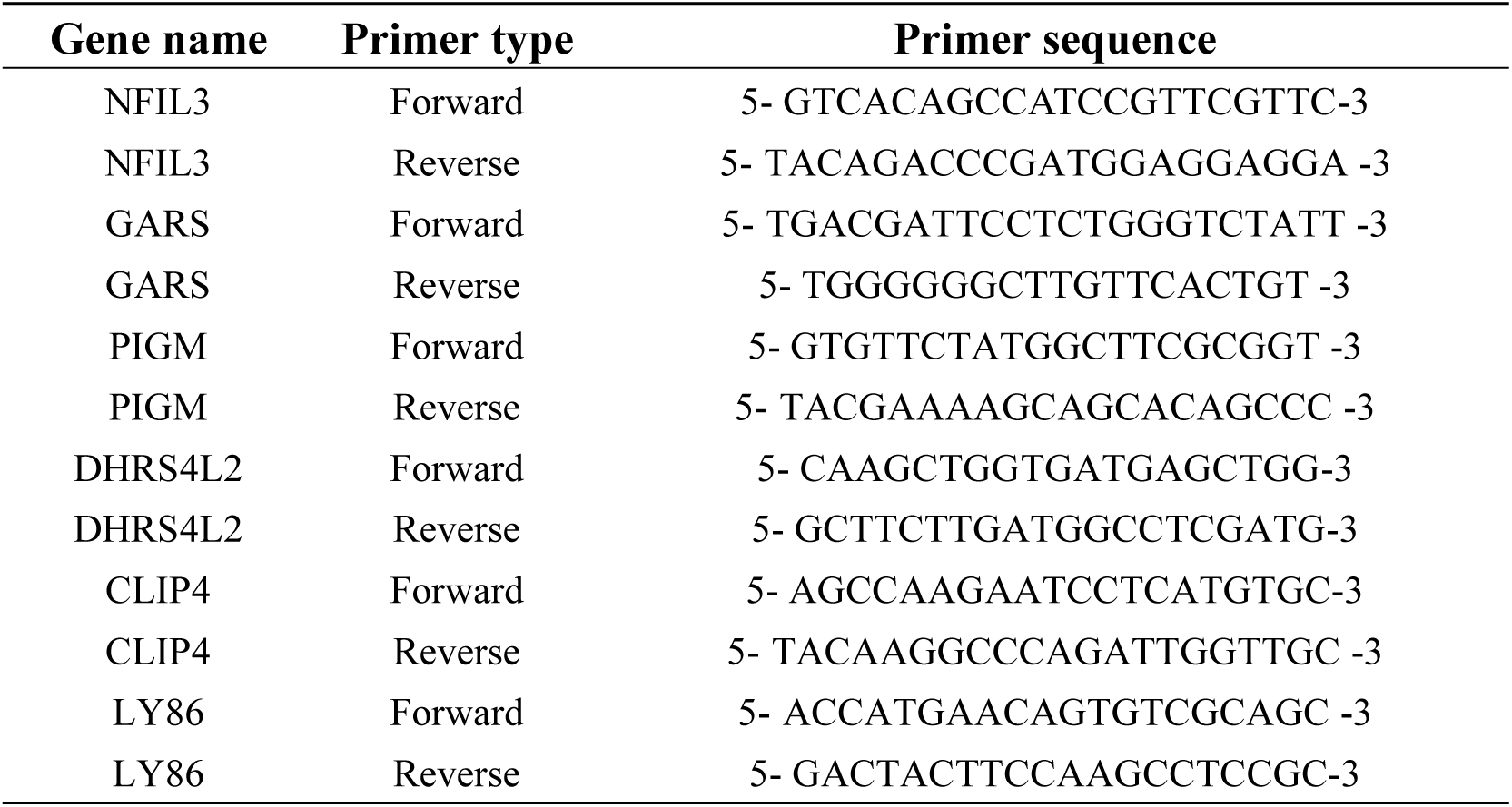
Primer sequences.

## 3. Results

### 3.1. Identification of DEGs

When the values for statistical significance were set as adjusted P-value < 0.05 and |log2FC|> 0.1, we identified 1671 DEGs, with 848 upregulated and 823 downregulated from the GSE66890 dataset, and 8964 DEGs from the GSE65682 dataset, with 4922 upregulated and 4042 downregulated, as represented in the volcano plots (Figures 1A,C), and top 20 DEGs that were simultaneously upregulated or downregulated in GSE66890 and GSE65682 were depicted in the heatmaps (Figures 1B,D).

**Figure 1.**
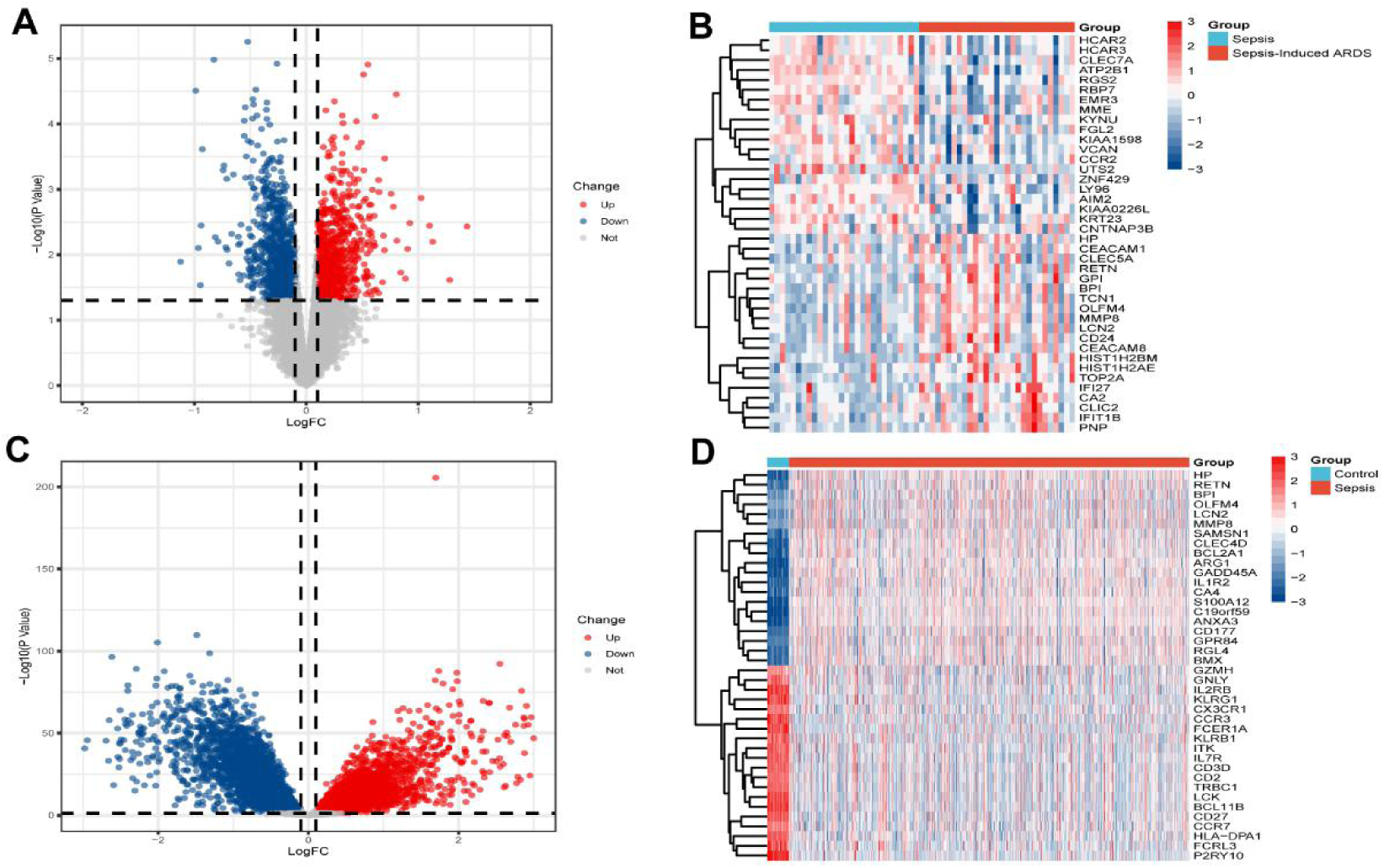
Identification of DEGs. (A) Volcano plot of DEGs between the Sepsis-induced ARDS group and the Sepsis group in GSE66890. (B) Heatmap displaying the top 20 upregulated and 20 downregulated DEGs in GSE66890. (C) Volcano plot of DEGs between the Sepsis group and the control group in GSE65682. (D) Heatmap displaying the top 20 upregulated and 20 downregulated DEGs in GSE65682.

### 3.2. The identification of modules related to Cellular Senescence through WGCNA

In our screening for cellular senescence-associated genes, mitochondrial energy metabolism served as an essential consideration, consistent with its established linkage to senescence^[16]^. Phenotype Cellular Senescence and Mitochondrial Energy Metabolism had strong correlation due to the heatmap (Figure 2A). The WGCNA algorithm was used to identify modules related to Cellular Senescence and Mitochondrial Energy Metabolism phenotype scores. When the scale-free topology fitting index reached 0.9, the soft-thresholding power b was 10 ((Figure 2B)). When the cutHeight = 150c, we obtained Sample Dendrogram and Trait Heatmap after dynamic tree cutting (Figures 2C). Under the parameter settings of minClusterSize =100 and mergeCutHeight=0.15, nine modules were identified (Figures 2D). The correlation between each module and disease status was subsequently analyzed. In the MEyellow module, Cellular Senescence, r = 0.86, P = 9.26e-18; Mitochondrial Energy Metabolism, r = 0.72, P = 2.13e-10; In the MEgreen module, Cellular Senescence, r = 0.85, P = 8.03e-17; Mitochondrial Energy Metabolism, r = 0.81, P = 2.91e-14, indicating that the MEyellow module was most significantly correlated with Cellular Senescence (Figures 2E). According to the correlation coefficient and P value, we selected the MEyellow module containing 556 genes as the key module for subsequent analysis.

**Figure 2.**
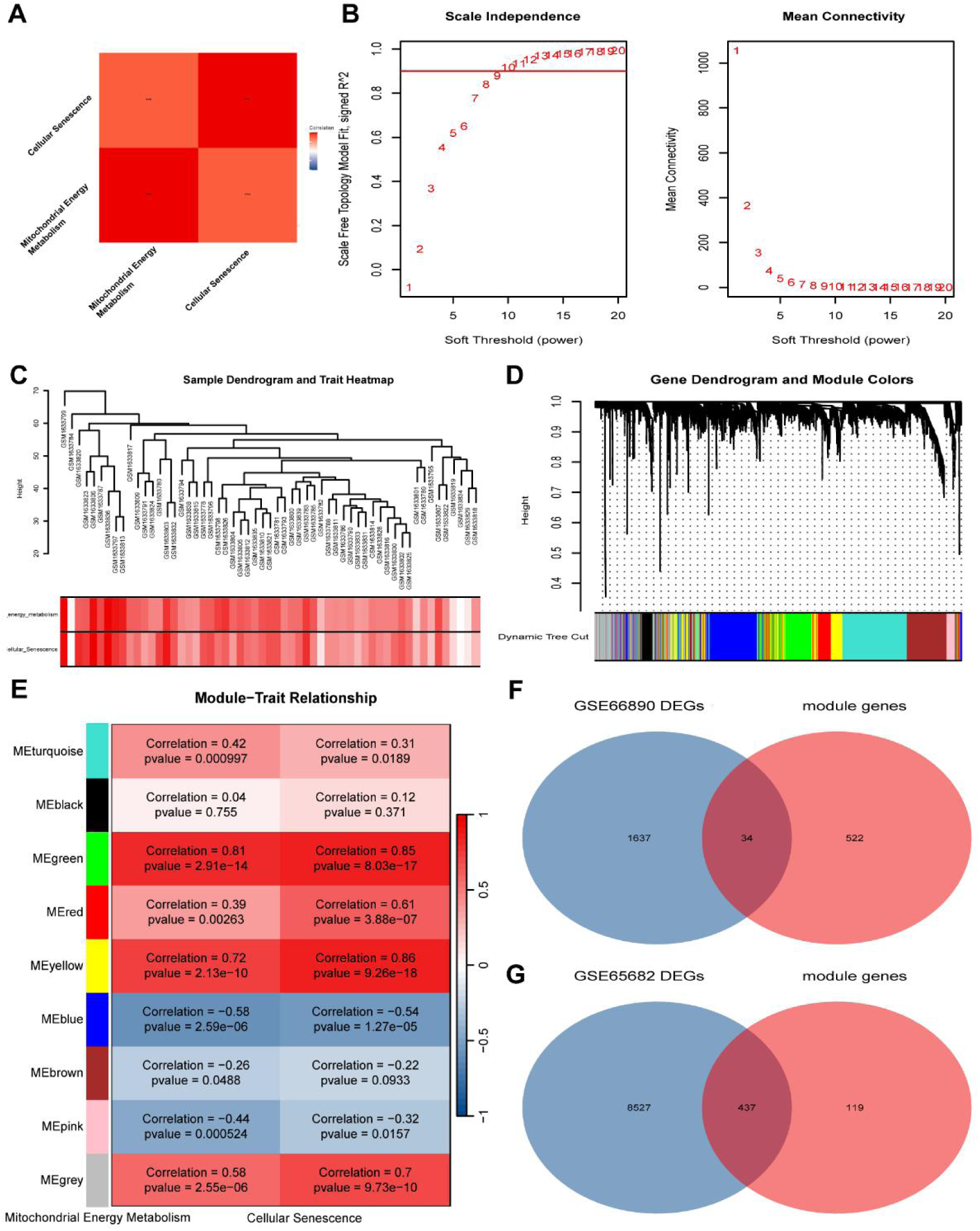
WGCNA identifying key gene modules associated with Cellular Senescence. (A) Phenotype scores correlation heatmap of Cellular Senescence and Mitochondrial Energy Metabolism. (B) Estimation of the value of the soft thresholding for a scale-free network of co-expression genes. (C)Sample Dendrogram and Trait Heatmap after dynamic tree cutting. (D) Gene dendrogram with identified modules via hierarchical clustering. (E) Association between modules and two phenotype related genes. (F) Venn diagram of GSE66890 DEGs and key module genes. (G) Venn diagram of GSE65682 DEGs and key module genes.

We further conducted intersection analyses between two datasets DEGs and key module genes to identify Sepsis and Sepsis-induced ARDS associated CS-DEGs, yielding 34 and 437 CS-DEGs respectively, with outcomes depicted in separate Venn diagrams (Figures 2F-G).

### 3.3. Screening and identification of characteristic Cellular Senescence biomarkers for Sepsis and Sepsis-Induced ARDS

To evaluate the diagnostic value of the 34 identified CS-DEGs in Sepsis-Induced ARDS, we performed a logistic regression analysis based on GSE66890 training dataset. The results demonstrated that 26 CS-DEGs were statistically significant (p < 0.05).

We further used six ML algorithms, including bagged trees, RF, Bayesian analysis, Boruta method, LASSO, and LVQ to analyze the relationship between the CS-DEGs from our logistic regression analysis and sepsis-induced ARDS event occurrence in the GSE66890 dataset.

Applying the bagged trees algorithm (Figure 3A), we identified 26 CS-DEGs that are all closely related to Sepsis-Induced ARDS event occurrence: XPNPEP1, CKAP5, FAM198B, PIGM, CTSO, GARS, LY86, MCM7, STRBP, DHRS4L2, LYZ, NFIL3, TNFSF13B, CLIP4, NOP16, HNMT, POGLUT1, CXCL16, IFNAR1, 1-Mar, AHR, MPEG1, PSMC5, CCR2, PLCB1, VCAN. Using the RF algorithm (Figure 3B), we screened 23 CS-DEGs closely associated with Sepsis-Induced ARDS-related events: PIGM, CKAP5, FAM198B, GARS, XPNPEP1, LYZ, LY86, STRBP, NFIL3, MCM7, CLIP4, DHRS4L2, POGLUT1, CXCL16, CTSO, IFNAR1, 1-Mar, CCR2, NOP16, HNMT, PSMC5, AHR, TNFSF13B, MPEG1. Using the Bayesian algorithm (Figure 3C), we identified 21 CS-DEGs that are closely related to Sepsis-Induced ARDS event occurrence: XPNPEP1, PIGM, FAM198B, NFIL3, GARS, CLIP4, CCR2, AHR, DHRS4L2, LY86, HNMT, MPEG1, CKAP5, MCM7, POGLUT1, NOP16, CTSO, 1-Mar, VCAN, IFNAR1, LYZ. Using the wrapper (Boruta) algorithm (Figure 3D), we selected 11 CS-DEGs closely connected to Sepsis-Induced ARDS event occurrence: STRBP, XPNPEP1, CKAP5, GARS, DHRS4L2, LYZ, NFIL3, PIGM, CLIP4, FAM198B, LY86. Employing the LASSO algorithm (Figure 3E), 10 CS-DEGs closely related to Sepsis-Induced ARDS event occurrence were extracted: CCR2, CLIP4, CTSO, CXCL16, DHRS4L2, GARS, HNMT, LY86, MCM7, NFIL3, NOP16, PIGM, POGLUT1, STRBP, XPNPEP1. Finally, using the LVQ algorithm (Figure 3F), we identified all 26 CS-DEGs closely related to Sepsis-Induced ARDS event occurrence.

**Figure 3.**
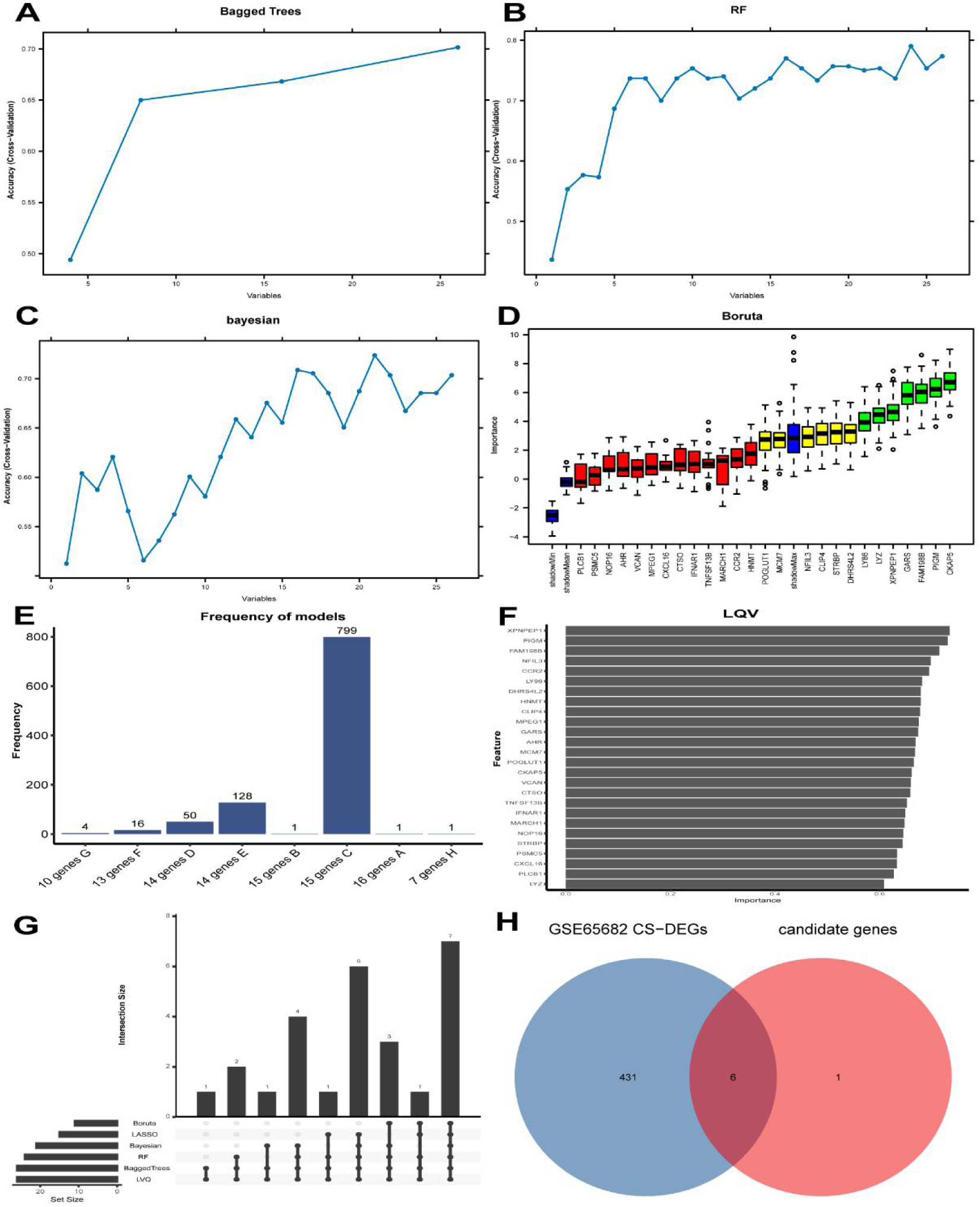
Differential Gene Expression and CS-DEGs Analysis using Machine Learning Algorithms. (A) Bagged trees algorithm value distribution for different variables and model precision. (B) Precision value distribution of the RF algorithm for different numbers of variables. (C) Bayesian algorithm precision value distribution with different numbers of variables. (D) Boruta algorithm. Genes in the green and yellow boxes were selected for inclusion in the model. (E) Frequency distribution of LASSO algorithm results over 1,000 repeated experiments. Bar chart values indicate the frequency of results for each gene set. (F) Importance values of key genes identified using the LVQ algorithm. (G) Visualization of the upset plot of six different ML algorithms. (H) Venn diagram of GSE65682 CS-DEGs and candidate genes.

Because different ML algorithms focus on different aspects, we retained the genes in all six ML algorithm results as candidate genes, resulting in a total of 7 genes (Figure 3G): CLIP4, DHRS4L2, GARS, LY86, NFIL3, PIGM and XPNPEP1 through the Upset analysis. Next, we performed intersection analysis of these candidates with CS-DEGs from the GSE65682 dataset (Figure 3H), and 6 final genes were selected as hub genes, which were NFIL3, GARS, PIGM, DHRS4L2, CLIP4, and LY86.

### 3.4. Evaluation of diagnostic performance and nomogram construction based on Hub genes

We further evaluated the diagnostic efficacy of hub genes as Cellular Senescence-related biomarkers.

In GSE66890 training set, the expression levels of NFIL3, GARS, PIGM, DHRS4L2, CLIP4, and LY86 were significantly different in the sepsis-induced ARDS group versus the sepsis group (Figure 4A). A nomogram model was constructed based on these six genes in the training set (Figure 4C), the AUC values was 0.89 (Figure 4D), confirming the potential application of this model in sepsis-induced ARDS diagnosis.

**Figure 4.**
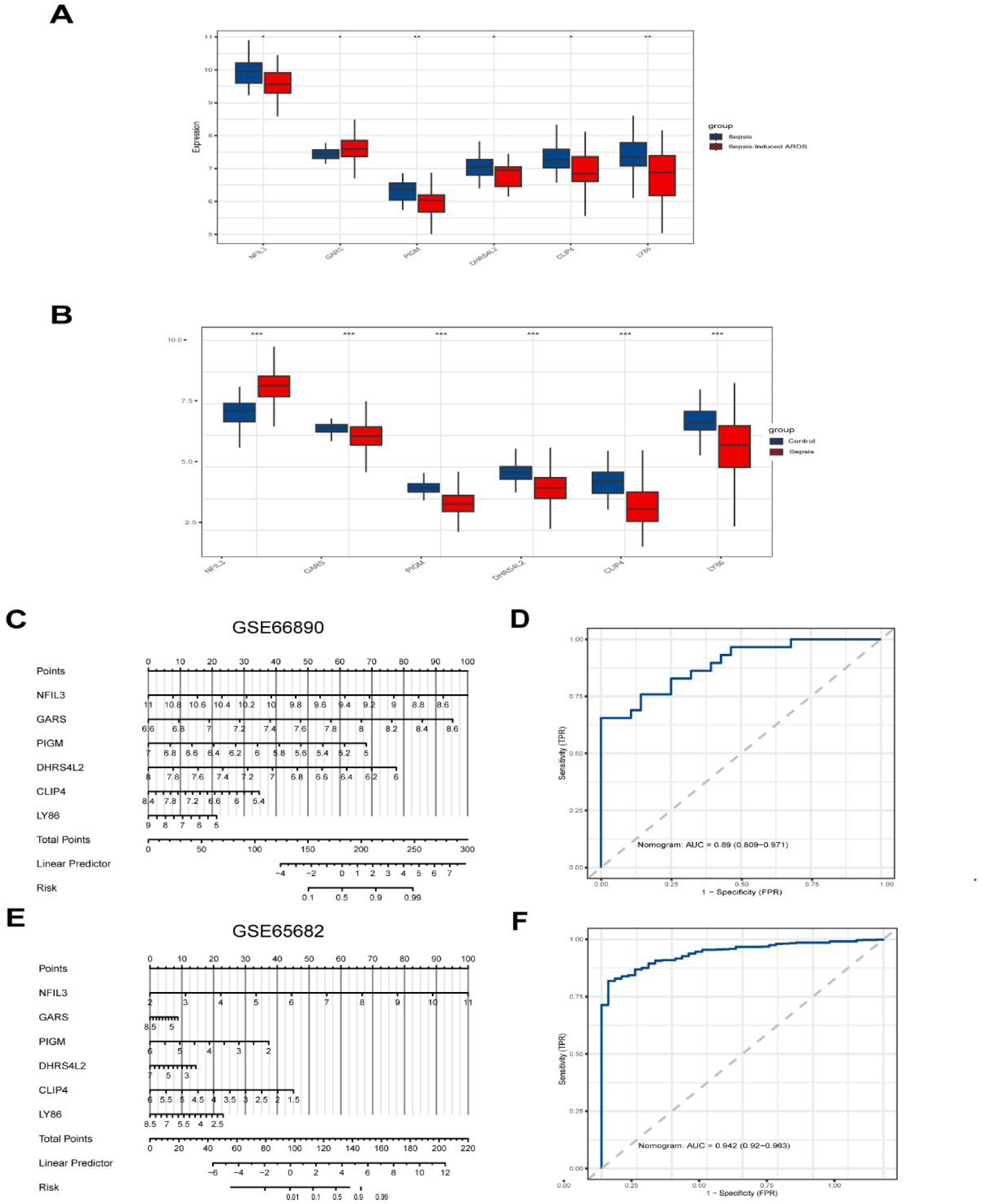
The expression levels of hub genes and Development of the diagnostic nomogram model: (A) The expression levels of hub genes in In GSE66890 training set. (B) The expression levels of hub genes in GSE65682 validation set. (C) Nomogram model based on the six genes in sepsis-induced ARDS. (D) ROC curve of the sepsis-induced ARDS diagnosis model. (E)Nomogram model based on the six genes in sepsis. (F) ROC curve of the sepsis diagnosis model.

In validation set, the expression levels of these six genes remained significantly different in the sepsis group compared to the control group (Figure 4B). The nomogram model (Figure 4E) based on these six genes achieving an AUC value of 0.942 (Figure 4F), which indicates that the hub genes had significant diagnostic potentiality.

### 3.5 Immune Infiltration Analysis

Based on ssGSEA algorithm, we assessed the infiltration levels of 28 immune cells in the different groups based on two datasets. Cumulative bar charts depicted the proportional representation of each immune cell subtype (Figures 5A-B). In training set, the bar chart revealing significant alterations in the proportions of activated CD56bright natural killer cell, Eosinophil, Macrophage, MDSC, Natural killer cell, and Neutrophil in the sepsis-induced ARDS group. Within the validation set, proportions of all immune cell subtypes except effector memory CD4 T cells were significantly altered in the sepsis group.

**Figure 5.**
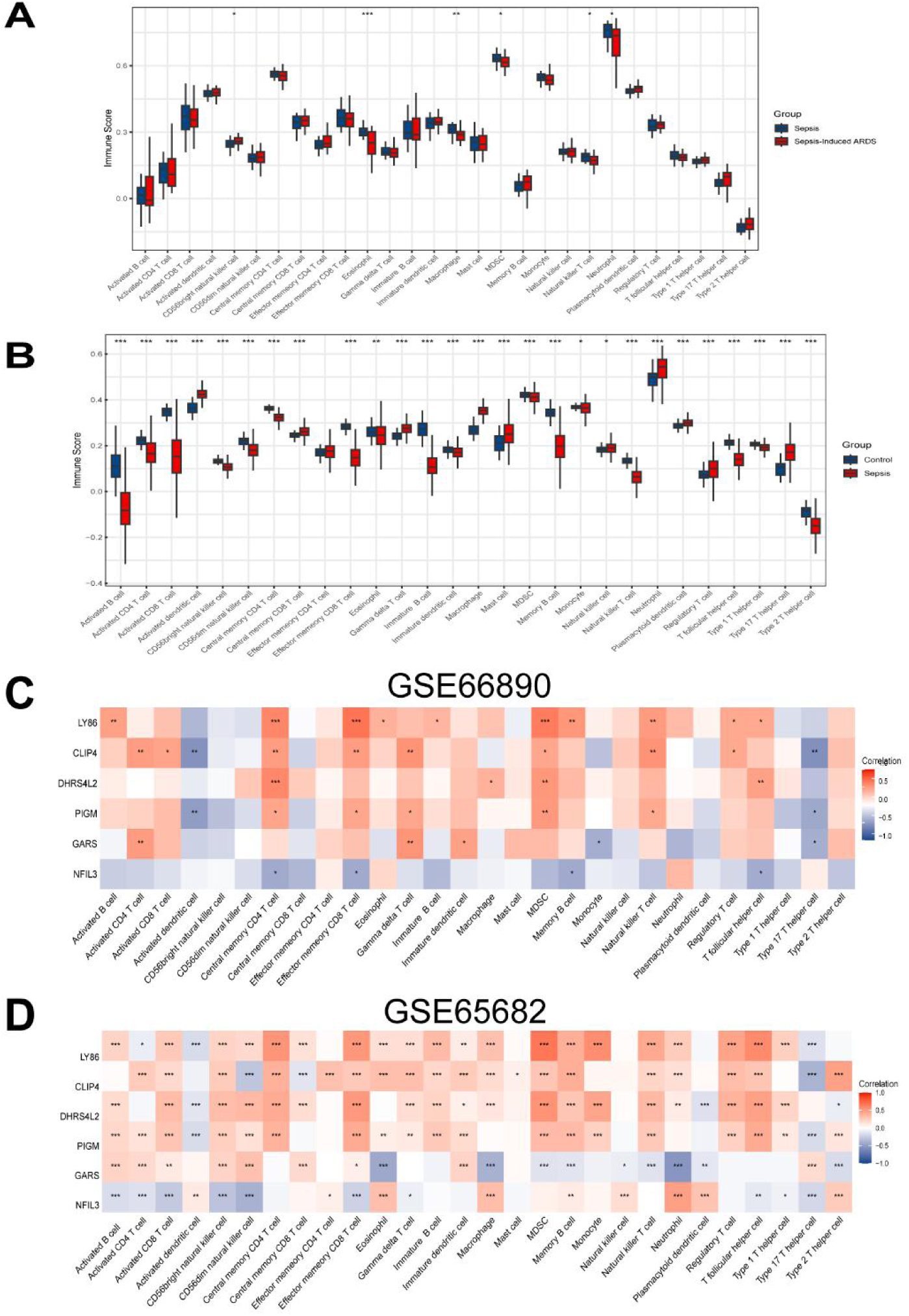
Immune infiltration analysis and correlation with hub genes in sepsis-induced ARDS. (A) Differential immune cell infiltration between sepsis and sepsis-induced ARDS groups. (B) Differential immune cell infiltration between Control and sepsis groups. (C-D) Correlation heatmap between hub genes and immune cell subtypes in two datasets.

Heatmap analysis identified significant correlations between immune cell subtypes and hub genes. For instance, in GSE66890, LY86 was strongly positively correlated with Central memory CD4 T cell, Effector memory CD8 T cell, and MDSC, whereas NFIL3 was negatively correlated with Central memory CD4 T cell, Effector memory CD8 T cell, Memory B cell, and T follicular helper cell (Figures 5C-D).

Additionally, we assessed the associations between Cellular Senescence-related biomarkers (NFIL3, GARS, PIGM, DHRS4L2, CLIP4, and LY86) and the immune cells based on spearman correlation analysis in training set. Immune cells exhibiting the strongest correlation with the single gene were extracted, and presented in the form of correlation scatter plots (Figures 6A-F).

**Figure 6.**
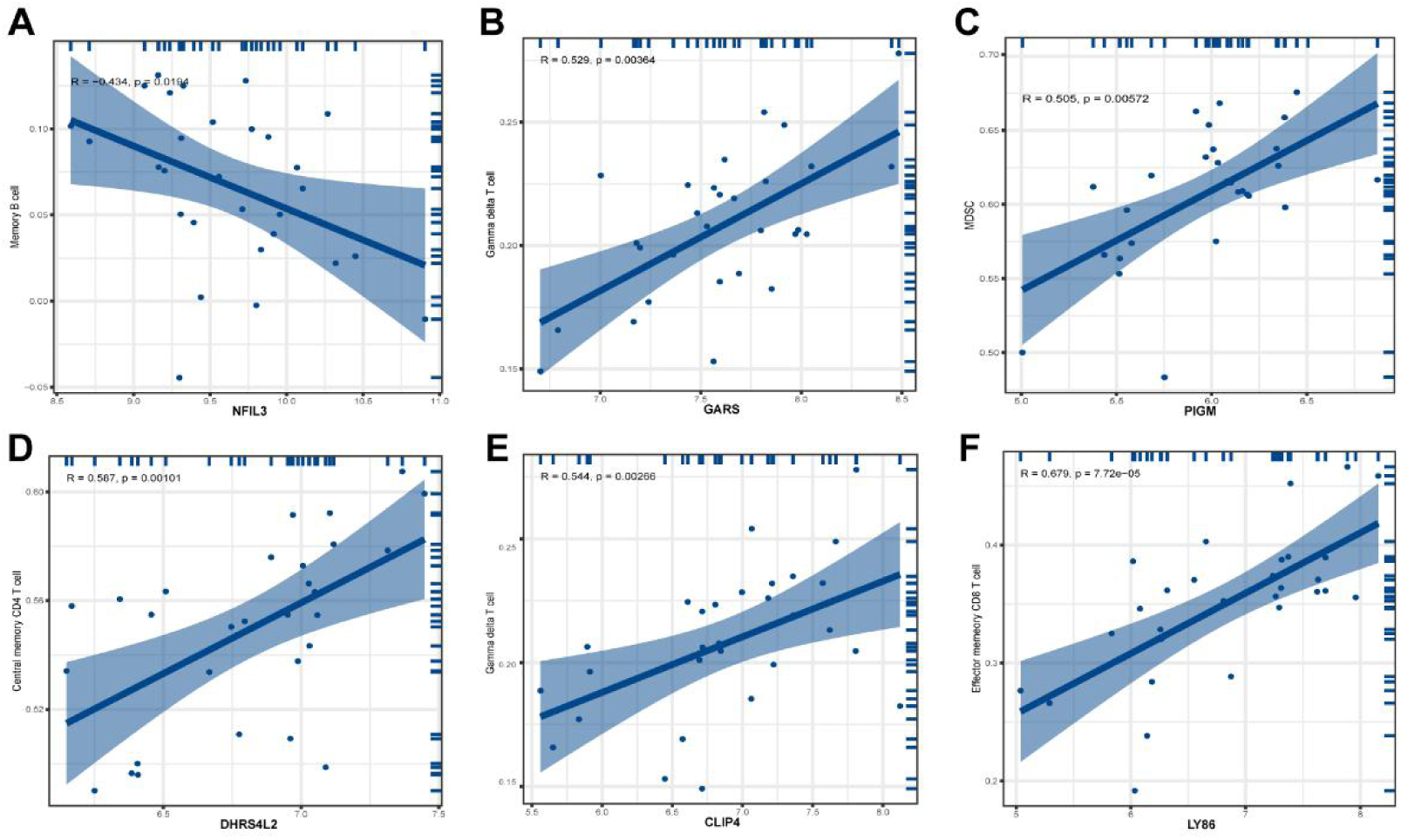
(A-F) Dot plots of correlation analyses between NFIL3 (A), GARS (B), PIGM (C), DHRS4L2 (D), CLIP4 (E), and LY86 (F) and immune cell types in dataset GSE66890.

These results suggest that Cellular Senescence may play a role in the development and progression of sepsis and sepsis-induced ARDS by modulating specific immune cell types within the immune microenvironment.

### 3.6 Identification of Cellular Senescence-related subtypes in Sepsis-Induced ARDS

To further elucidate the underlying mechanisms of Cellular Senescence in sepsis-induced ARDS, consensus clustering analysis was performed on the sepsis-induced ARDS samples. According to the cumulative distribution function (CDF) and delta area curve of the consensus matrix, the optimal cluster number was determined as k = 2 (Figures 7A-C). Therefore, the sepsis-induced ARDS samples were divided into two subgroups (Cluster 1: 14 samples; Cluster 2: 15 samples).

**Figure 7.**
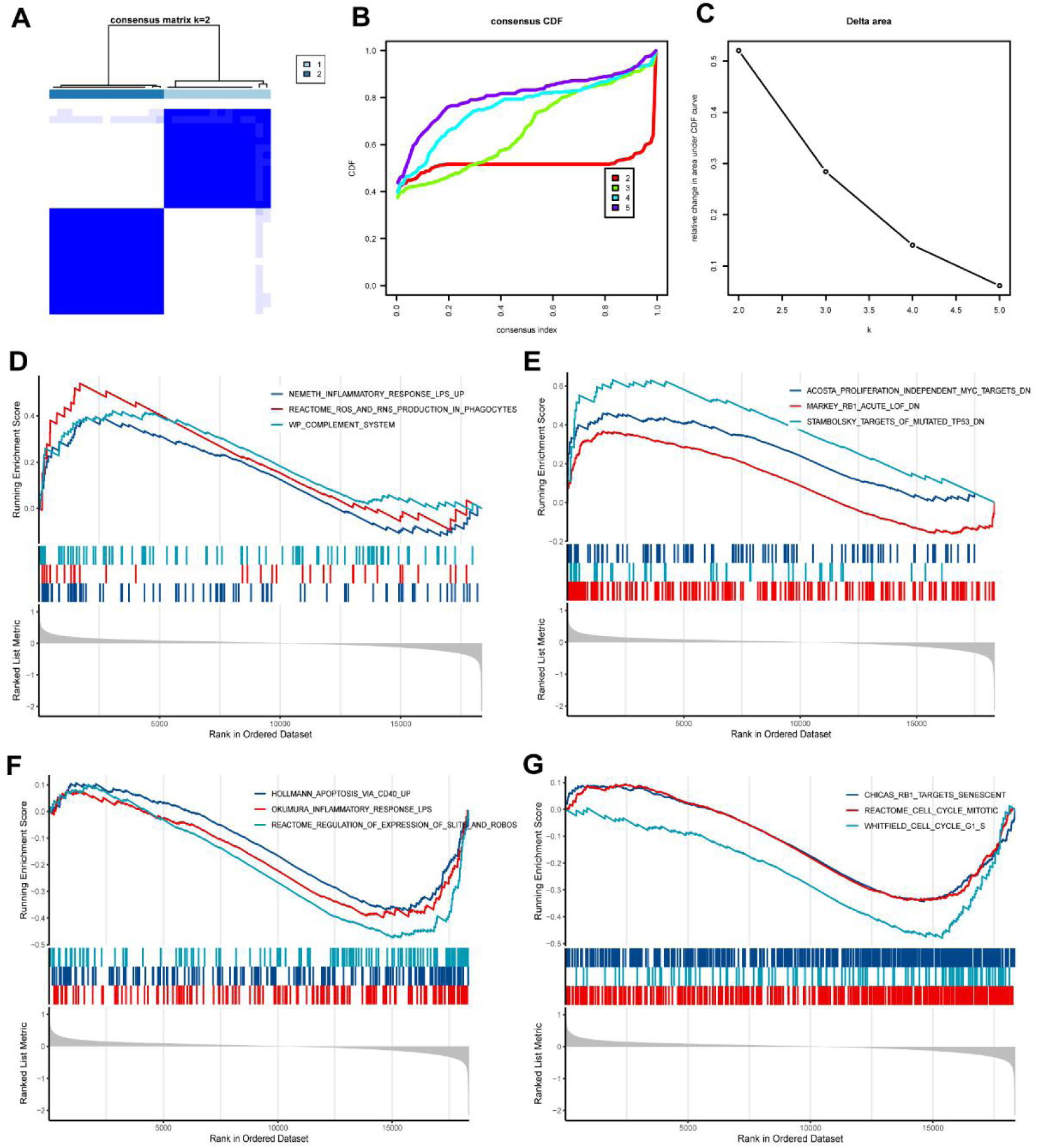
Analysis of Cellular Senescence -related molecular patterns in sepsis-induced ARDS. (A) Consensus clustering matrix at k = 2. (B) CDF curves for k values 2–5. (C) CDF delta area curve showing optimal clustering at k = 2. (D-G) GSEA pathways enriched in sepsis-induced ARDS.

In addition, GSEA was applied to explore the functional signatures in sepsis-induced ARDS based on two subgroups. The upregulations signaling pathways and the downregulation pathways which were significantly enriched were showed as Figures 7D-G. The above biological processes were closely related to the immune system and cell cycle.

### 3.7 Single cell analysis

By ‘FindallMarkers’ function, significant DEGs in every cluster were identified and then cell types were identified in an unbiased manner by the specific markers. Using UMAP analysis, we observed that six cell clusters were identified, including B cells, Monocytes, Neutrophils, NK cells, Platelets, T cells (Figure 8A).

**Figure 8.**
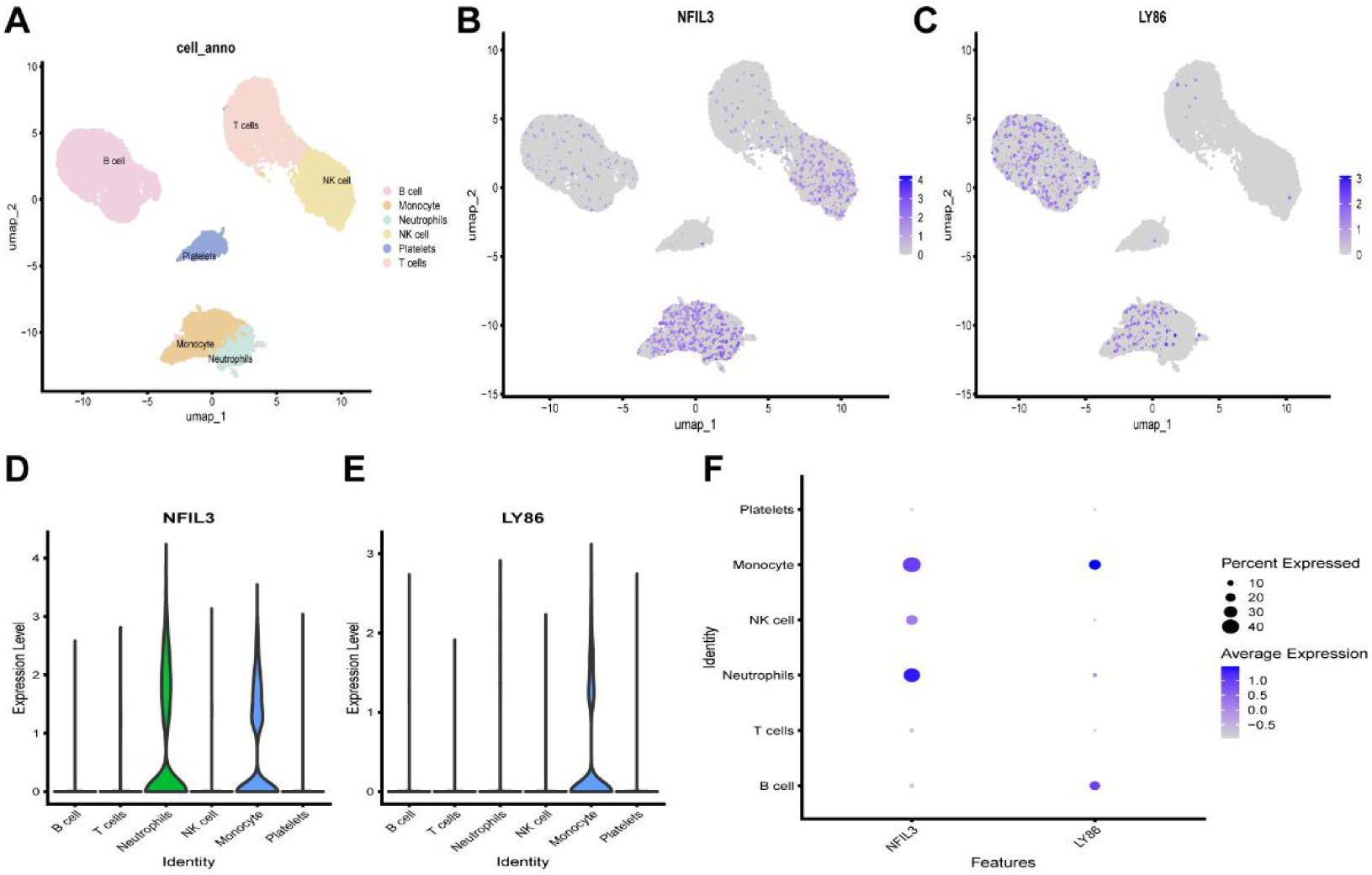
Single cell sequencing analysis of GSE167363 and cell localization of NFIL3 and LY86. (A) Classification of cells and a total of 6 cell types can be found. (B-F) Expression of 2 hub genes in single cells.

We assessed hub genes expressions in distinct cell classes, two genes (NFIL3 and LY86) showed significantly differential expression, and the cell classes involved in the higher expression of the hub genes were screened out. Subsequently, two hub genes expressions in each cell type were visualized respectively by feature Plots, violin plots and dot plots (Figures 8B-F). The expression of NFIL3 and LY86 were both higher in Monocytes, while NFIL3 was also higher in Neutrophils.

### 3.7 Validation of Hub Genes Expression

RT-qPCR analysis confirmed significant differential expression of NFIL3 in LPS-stimulated dHL-60 neutrophil-like cells compared to the control group (Figure 9), consistent with our bioinformatics predictions. Therefore, it is important to further explore NFIL3 gene in sepsis-induced ARDS.

**Figure 9.**
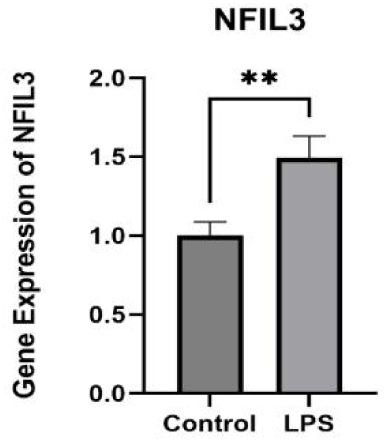
Expression of the NFIL3 gene in Control group and LPS group. **p < 0.01.

## 4. Discussion

Sepsis often induces multi-organ dysfunction as a result of a dysregulated host immune response, a particularly prevalent and severe pulmonary complication is sepsis-induced ARDS, which manifests as an intense pulmonary inflammatory storm marked by uncontrolled cytokine release and significant oxidative stress^[17]^. Since effective clinical management of sepsis-induced ARDS relies heavily on early prediction and diagnosis, bioinformatic analysis of large-scale gene expression data represents a powerful strategy for uncovering its molecular mechanisms^[18]^.

Accumulating evidence indicates that cellular senescence plays a critical role in the pathogenesis of sepsis-induced ARDS. Acting as a double-edged sword, cellular senescence can exert both protective and detrimental effects in pulmonary disorders. For instance, mesenchymal stem cells (MSCs) promote a shift toward senescent neutrophils in acute lung injury^[19]^, indicating a potentially beneficial role of induced senescence in this setting. Conversely, senescence in lung endothelial cells disrupts cell–cell junctions, compromises barrier integrity, and facilitates neutrophil adhesion and migration, thereby promoting age-related pulmonary inflammation and associated pathology^[20]^. Thus, a comprehensive identification of key mediators and signaling pathways associated with cellular senescence in sepsis-induced ARDS through bioinformatic approaches will clarify its exact role in disease mechanisms.

Mitochondria undergo substantial remodeling during cellular senescence, characterized notably by an increase in their size. This enlargement results from the accumulation of dysfunctional mitochondria, which generate excessive ROS ^[21]^. The persistence of such impaired mitochondria has deleterious consequences, disrupting cellular homeostasis and promoting degenerative changes in tissues^[16, 22]^. In light of the well-established connection between mitochondrial energy metabolism and cellular senescence, we specifically focused on cellular senescence-associated genes involved in mitochondrial energy metabolism. Our correlation heatmap visually confirms a strong positive association between these two phenotypic domains.

In our study, six hub genes were identified, NFIL3, GARS, PIGM, DHRS4L2, CLIP4, and LY86. These genes may play critical roles in the progression of sepsis-induced ARDS. NFIL3, a basic leucine zipper transcription factor, modulates circadian rhythms, neurodevelopment, and immune regulation in both innate and adaptive immunity^[23, 24]^. In our single-cell analysis, NFIL3 expression was elevated in monocytes and neutrophils. RT-qPCR further confirmed its significant differential expression, suggesting a key role in sepsis pathogenesis. Notably, inhibition of NFIL3 has been shown to alleviate sepsis-associated acute kidney injury (SA-AKI) by suppressing ACSL4-regulated ferroptosis and inflammation, highlighting the involvement of this rhythm gene in SA-AKI^[25]^. Thus, we speculate that NFIL3 may contribute to sepsis-induced ARDS through a similar mechanism, which warrants further comprehensive investigation. Glycyl-tRNA synthetase (GARS), a bifunctional member of the aminoacyl-tRNA synthetase (ARS) family localized in both cytoplasmic and mitochondrial compartments^[26]^, promotes oncogenic processes in breast cancer by accelerating cell growth, cell-cycle progression, colony formation, migration, and invasion via mTOR signaling^[27]^. Additionally, GARS enhances the infiltration of multiple immune cells, and high GARS levels in hepatocellular carcinoma (HCC) tissues correlate with increased immune cell infiltration^[28]^. These findings imply that GARS may participate in sepsis progression by regulating the cell cycle and modulating immune infiltration. PIGM plays important roles in multiple biological processes, including cell proliferation, differentiation, and apoptosis^[29]^. It encodes phosphatidylinositol glycan anchor biosynthesis class M, an enzyme involved in the phosphatidylinositol signaling pathway^[30]^. Notably, PIGM expression exhibits a negative correlation with the proportions of CD8⁺ T cells and CD4⁺ T cells ^[29]^. suggesting its potential functional relevance in sepsis. DHRS4L2 encodes a retinol oxidoreductase implicated in retinol homeostasis and is transcriptionally regulated by DNA methylation^[31]^. This gene is frequently downregulated in both leukemia and solid tumors ^[32]^. However, its role in sepsis remains to be elucidated. The CAP-Gly domain-containing linker protein family member 4 (CLIP4), a member of the CLIP-170 family, links organelles to microtubules via its CAP-Gly domain. This domain is crucial for protein–protein interactions, particularly in cytoskeletal organization and cell migration^[33]^. While CLIP4 has been reported to perform diverse functions across different cancer types^[34]^, its role in sepsis remains unclear. Lymphocyte antigen 86 (LY86, also known as MD-1) encodes a secreted glycoprotein involved in inflammation, humoral immunity, apoptosis, and signal transduction^[35, 36]^. Although the function of LY86 in sepsis has not been fully defined, our study observed elevated expression of LY86 in monocytes, providing preliminary insight into its potential mechanisms.

Utilizing these hub genes, we constructed a diagnostic model in the training set. The model showed high predictive accuracy for sepsis-induced ARDS. Conventional sepsis treatments, such as fluid resuscitation and broad-spectrum antibiotics, offer limited efficacy and are associated with notable drawbacks, including fluid overload, antibiotic resistance, and disruption of the gut microbiota^[37, 38]^. In contrast, bioinformatic approaches enable the prediction of sepsis onset and progression through gene-expression profiling^[39]^, providing a pathway toward precision medicine.

In the pathogenesis of sepsis and sepsis-induced ARDS, immune dysregulation plays a central role, with numerous immune cell populations actively participating in this process^[40, 41]^. In this study, we performed ssGSEA-based immune infiltration profiling to unravel the underlying immunological mechanisms. In the training set, comparative immune infiltration analysis revealed a significant reduction in five immune cell subsets, eosinophils, macrophages, MDSCs, natural killer cells, and neutrophils, in sepsis-induced ARDS compared to sepsis controls, along with elevated infiltration of CD56bright natural killer cells. These findings underscore immune dysfunction as a critical pathophysiological link between ARDS and sepsis.

In addition, we stratified patients into two molecular subtypes, High-aging and Low-aging, based on hub gene expression. GSEA revealed significant enrichment of cellular senescence-related genes in the Toll-like receptor 4 (TLR4) signaling pathway. It has been reported that baseline TNF-α production by circulating leukocytes is elevated, while responses to TLR2 and TLR4 agonists are impaired with aging, which may increase susceptibility to sepsis in elderly populations^[42]^.

While the findings of this study are promising, several limitations must be acknowledged. The analysis was based solely on publicly available datasets from the GEO database, and the number of datasets included was limited. Future research should integrate transcriptomic or single-cell sequencing data obtained directly from clinical samples of patients with sepsis-induced ARDS to improve the robustness and generalizability of the findings. Although animal models provided preliminary validation, clinical studies remain crucial to confirm the diagnostic and prognostic value of the identified hub genes in real-world clinical contexts.

## Conclusion

In summary, this study identified six hub genes linked to cellular senescence (NFIL3, GARS, PIGM, DHRS4L2, CLIP4, and LY86) in sepsis-induced ARDS using integrated bioinformatics. Diagnostic and prognostic models based on these genes effectively distinguished sepsis patients with ARDS. Immune infiltration and single-cell analysis connected these genes to monocytes and neutrophils, implicating them in immune and cell cycle pathways. NFIL3 was validated as upregulated in septic rat lungs, highlighting its potential as a biomarker and therapeutic target.

## Author contributions

**Panpan Li:** Data curation, Formal analysis, Methodology, Project administration, Software, Validation, Visualization, Writing – original draft; **Yongbing Yu:** Formal analysis, Investigation, Methodology, Validation, Writing – original draft; **Jihua Feng:** Funding acquisition, Investigation, Resources, Supervision, Writing – review & editing; **Shanshan Huang:** Formal analysis, Investigation, Software, Visualization; **Jianfeng Zhang:** Conceptualization, Funding acquisition, Methodology, Project administration, Resources, Writing – review & editing. All authors agree to be accountable for all aspects of the work.

## Acknowledgements

Thanks to the members of our laboratory for their contributions.

## Data availability statement

All the datasets could be downloaded directly from the indicated websites.

